# Objective comparison of 3D dental scans in forensic odontology identification

**DOI:** 10.1101/2025.03.31.25324929

**Authors:** Anika Kofod Petersen, Dorthe Arenholt Bindslev, Andrew Forgie, Palle Villesen, Line Staun Larsen

## Abstract

In forensic odontology disaster victim identification, it is crucial to assess the similarity between *post mortem* (PM) dentitions and *ante mortem* (AM) dental records from a database. To facilitate ranking AM records by likelihood of a match, the similarity evaluation must yield an intuitive, quantitative score. This study introduces a scoring scheme designed to effectively distinguish 3D dentition surfaces acquired by intraoral 3D photo scans. The scoring scheme was validated on an independent dataset. The scoring scheme presented utilizes two levels of surface similarity, spanning from local similarity of surface representations to regional similarity based on relative keypoint placement. The scoring scheme demonstrated exceptional discriminatory power on the validation data, achieving a Receiver Operating Characteristic (ROC) area-under-the-curve (AUC) of 0.990 (95% CI 0.988 to 0.992). Implementing such a scoring system in disaster victim identification workflows, where AM 3D data can be procured, can provide an initial likelihood of matching, enabling forensic professionals to prioritize cases and allocate resources more efficiently based on objective measures of dental similarity.

## Introduction

In the aftermath of disasters, the accurate identification of victims is essential. INTERPOL has categorised three primary identifiers, deemed as the most accurate methods for identification purposes: DNA, fingerprints, and dental comparison [1,2]. With, clinical dental records, it is possible to build disaster specific dental databases, making dental comparison a highly useful identification method in many disasters [1]. With the rise of use of intraoral 3D surface scanners in dentistry, many dental records contain digital 3D dental surface models that can prove useful in identification cases, especially when the deceased have had little dental work done, leaving fewer details in relation to the use of common 2D data [1,3,4]. As to our knowledge, there is currently no consensus on how to use these 3D dentition surface models for identification purposes, even though they might aid in the automation of parts of the disaster victim identification (DVI) process [4,5].

To automate parts of the DVI process, the *ante mortem* (AM) 3D dentition surface models could be compared to *post mortem* (PM) intraoral 3D surface scans (IOS) in order to give a quantitative score of matching potential [5]. Prospectively, such a quantitative score could be used to score the similarity of the PM dentition surface to an AM database, thus giving a preliminary indication of matching, or a ranking of the AM database for the forensic odontologist to use [5].

Few methods with a quantitative score indicating matching status have been proposed for automatic 3D dental comparison of AM-PM IOS [6-10]. Conventional 3D matching by superimposition does not account for the variation within dentitions across time and trauma, e.g. the natural mesial drift of teeth and partial dentitions caused by mechanical trauma during a disaster [6]. Such superimposition methods work on a global surface area, meaning that the sum of dentition surfaces (the complete dentition considered as a whole) contribute to the scoring, potentially risking intrapersonal variation between time points to discard a correct match [6,11-13].

A solution could be to not use the entire dental surface of each tooth but use representative measures. Zhong and colleagues used keypoints representing the dental arch which spans the global dental surface [8]. But, when using the dental arch, one must consider that 3D scans (IOS) are known to be less accurate when spanning long distances, making the curve of the dental arch a poor measure of dental similarity to be used as the sole source of matching [14,15]

Another approach is to look at regional similarity, e.g. by focussing on molar surfaces or curvature signatures. This allows some amount of intrapersonal variation (individual changes over time due to tear and drift) and can still capture larger interpersonal variations [7,9,10,16].

Lastly, local similarity captures the immediate surface similarity within a limited range, spanning no more than a couple of teeth, thus allowing for large intrapersonal variations caused either naturally or by the disaster conditions [5,16]. To minimise the impact of intrapersonal variation one could use specific points of interest on the teeth, termed keypoints [7,9]. Zhang and colleagues compared dentition surfaces using a custom representation method, Signed Feature Histogram (SFH), but did not attach keypoints to extreme curvatures [7]. The keypoints used by Zhang et. al were found by a Random Forest model which has not been publicly deposited, thus limiting the proper use of SFH to manually annotated dental surfaces [7]. SFH was tested using keypoints found at curvature extremes, but it was outperformed by SHOT representation [5]. Zhou and colleagues combined the extreme curvature keypoints approach and the surface representation approach [9]. This was done by downsampling the surface point clouds in accordance with extreme surface curvatures prior to keypoint representation by Fast Point Feature Histogram (FPFH) [9]. By reducing the point clouds with about 92% prior to representation, the keypoint representations were forced to only include the relative position of keypoints and did therefore not truly include the local surface curvatures, but rather the regional extreme curvature landscape.

In addition, we tested fifteen different approaches for keypoint descriptions of dental surfaces and showed that Difference of Curvature (DoC) keypoint detection and Signed Histogram of OrienTations (SHOT) keypoint representation showed promising results for automatic 3D dental comparison [5]. These previous promising findings were based on dental data with imputed difference, meaning that the noise observed in that data was not guaranteed to be reflective of the noise to be observed on the dentition surface due to dental changes happening in a living individual during the passing of time [5].

In this study, we aim at testing and improving the keypoint pipeline previously proposed, using data with true difference caused by the passing of time.

## Materials and Methods

### Data

To investigate and validate a quantitative scoring methodology for the keypoint pipeline, without biasing the conclusion of performance, we needed two independent datasets: one to develop the scoring scheme, and another to test the validity of the scoring method.

For development, we were granted permission to collect a dataset from Tandplejen Silkeborg (Silkeborg Municipality Dental Care Facility) in Denmark, that consists of IOS from jaws of 1999 young individuals who have had their dentition scanned at the facility in relation to orthodontic planning/treatment. The data was filtered to only include IOS of jaws that were scanned two or more times with a time difference of more than 6 months (183 days), resulting in 404 IOS from 97 individuals. This data is referred to as the Silkeborg Municipality Dataset (SMD). The original SMD contained a total of 404 IOSs with the largest time difference being 8 years and a median time difference of 733 days. To include partial dentitions in the analysis, the IOS were copied and split computationally as follows: Each IOS was used in full and in two halves split by the sagittal plane and two halves split by the frontal plane (determined by the centre of mass within the scan). This gives a total of 5 versions of each IOS. This resulted in a total of 2020 IOSs in SMD. Since the IOS were part of dental treatment/planning, and the minimal time difference was 6 months, all IOS were expected to have clinically relevant changes to the dentition surface, including correction of misalignments, shedding of primary teeth and eruption of permanent teeth or even extraction of permanent teeth - which would make perfect matching impossible. We expected that it would be impossible to unambiguously differentiate SMD into matches and mismatches. For SMD, soft tissue was limited in the IOS by using grid-cutting [17]. In summary, we expected the SMD to cover large variation caused mainly by natural growth due to age and time span and tooth movement caused by orthodontic treatment, thus exceeding the variation to be expected in most real-world cases.

To capture the variation of IOS caused by different scanning times within reason of what is to be expected in a disaster scenario, we collected a dataset from recruited healthy adult volunteers, which will be referred to as the Healthy Volunteer Dataset (HVD).

A total of 51 healthy volunteers gave informed consent to participate in the HVD and were subject to two IOS scans with a median time difference of 6 months. The participants were between the ages of 23 to 61 years old, 23 were self-reported females and 28 were self-reported males.

To mimic partial dentitions the jaws were computationally split in half by the sagittal plane and the frontal plane, determined by the centre of mass within the scan after copying. Therefore, this dataset consists of 51 individuals with two jaws, scanned at two timepoints, with 5 versions of each IOS. That is a total of 1020 dental meshes to be compared by pairwise comparison. When comparing the IOS from HVD (all-vs-all), excluding identical pairs and partial identical pairs, and restricting by only comparing jaws of the same type (upper jaws vs. upper jaws and lower jaws vs. lower jaws), a total of 515,100 pairs were compared with 4,284 pairs being from the same individual (matches).

We expect this dataset to closely mimic the data that would be collected in a disaster scenario. For data preprocessing the dental scans were subject to grid-cutting in order to limit the amount of soft tissue, since oral soft tissue is expected to be affected by disaster conditions to a higher degree than dental hard tissue [17].

The HVD and the SMD were kept separate to avoid bias when evaluating method performance. During pairwise comparisons, all identical pairs (i.e. IOS-A compared with IOS-A without any augmentation) have been excluded, since such pairs gave a perfect match on all accounts.

### Comparing teeth by local similarity only

Briefly, the previously proposed pipeline has four steps: 1) Mesh preprocessing by grid-cutting [17], 2) keypoint detection by DoC [5,16], 3) Keypoint representation by SHOT [5] and 4) keypoint similarity scoring. With this setup, the previously proposed keypoint pipeline compares a pair of IOS (IOS-A and IOS-B) by measuring the Euclidean (L2) distance between all keypoints from IOS-A to IOS-B [5]. The distances are then sorted for each IOS-A keypoint to find the ratio between the two shortest distances for each IOS-A keypoint. As a result, each keypoint of IOS-A returns an L2 ratio. The distribution of L2 ratios for a given pair of scans should then reveal whether IOS-A and IOS-B originated from the same individual (a matching pair) or from different individuals (mismatching pair) [5].

Previously we presented the Inter Quartile Range (IQR) score to distinguish between matches and mismatches from the L2 ratio distributions [5]. Unfortunately, if the dentition has been subject to significant variation, a matching pair will have some keypoints from the first scan that no longer have a match in the second. In these cases, the L2 ratio distribution of matches from different sampling times would be a combination of the identical match and the mismatch distributions, causing the IQR score to fail in match/mismatch separation [5].

It is expected that some keypoints, by chance and due to the interpersonal similarity of dental surfaces, would score low L2 ratios, while others would score high L2 ratios due to intrapersonal dental changes [13,18]. Taking these assumptions into account, not all L2 ratios should be considered in a matching score. Therefore, this study differs from the previously proposed pipeline, by trying to optimise a scoring scheme used to identify matches.

For the new scoring system, we need to only include a subset of the keypoints, namely the subset that is expected to match between two IOS from different timepoints.

By only including the lowest L2 ratios from a comparison, we compare the keypoints most likely to have a true match. Then, the mean of L2 distances for this subset would represent how well the two IOS are matching. In this study we tested three types of means: arithmetic means, geometric means and harmonic means. Furthermore, the L2 distance ratio at the optimised index of the best performing method (arithmetic mean) was explored to investigate the effect of false positive matching keypoints. To choose how many keypoints to include, we optimised the differentiability of matches and mismatches in our SMD dataset, which were expected to have a high overlap between matches and mismatches. This type of similarity can be described as local similarity, since only the information within an immediate vicinity of a keypoint is included (Figure 1a).

**Figure 1.**
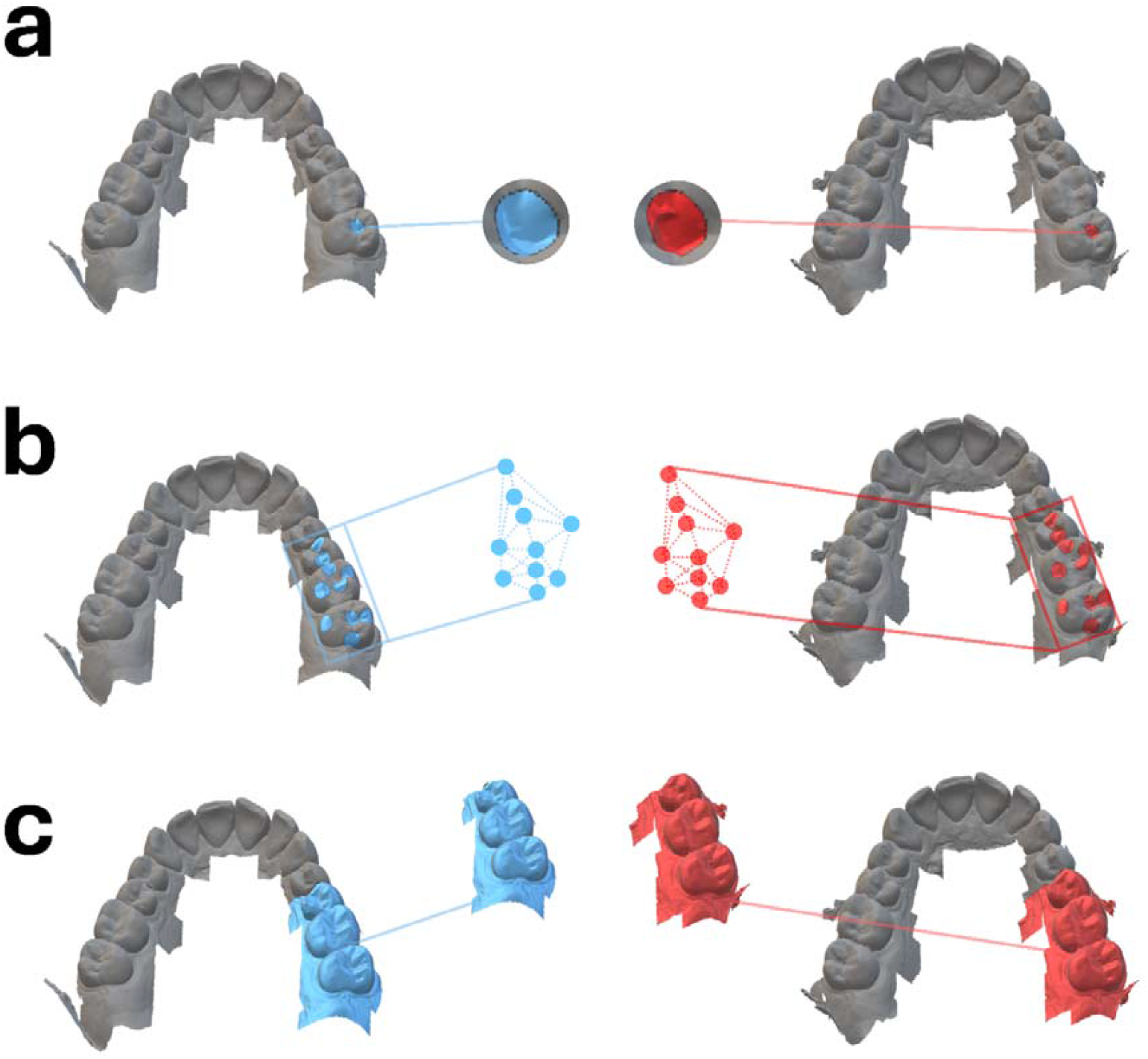
Types of similarity measures. Shown is a) local similarity, b) regional similarity and c) global similarity.

### Adding placement to local similarity

L2 distance means does not take keypoint placement into account, meaning that if a keypoint sitting on a molar in IOS-A has the best matching keypoint on an incisor in IOS-B, it is still considered a matching pair of keypoints. To further enhance the scoring method, how similar the keypoints are placed in relation to each other on the compared IOS can be considered. This is called the relative placement of keypoints, and describes the regional similarity (Figure 1b), since information spanning a region of the dentition surface is included. Again, by optimizing the differentiability of SMD, the relative placement of 8 keypoints was chosen to supplement the mean distances as the quantitative score. To score how similar the relative placement of the best matching 8 keypoints between IOS-A and IOS-B are, they were subject to a generalized Procrustes analysis (GPA). The disparity of the GPA is a measurement of how similar the relative placement of the keypoints is after minimizing differences caused by translation, rotation and scaling [19]. We now have four scores indicating similarity: arithmetic mean, geometric mean, harmonic mean and the GPA disparity. These four scores represent two types of similarity scores, local and regional, as described in Figure 1a-b. A combination of scores could have the advantage of taking both keypoint position (‘place-in-space’) and keypoint surface (‘vicinity curvature’) similarity into consideration. Therefore, we also tested using a weighted average of the two types of scores.

To compare the predicative power between the different scoring schemes, one must take into consideration the imbalance of the size of the two classes (matches and mismatches) in the dataset. The mismatch class is orders of magnitude larger than the matching class, which can cause bias when investigating performance. To alleviate this imbalance, HVD was subjected to stratified bootstrapping with random under-sampling of the majority class [20,21] prior to receiver operating characteristic (ROC) area under the curve (AUC) [20,22] and precession-recall (PRC) AUC investigation [23]. This was done for a total of 100 bootstraps, and the confidence interval of the AUC was calculated based on the aggregated AUC for each bootstrap.

## Results

### Estimating a good model using the SMD data

To decide how many keypoints to be used for a matching score, we aimed at optimising the separation of matches and mismatches within the SMD. To validate that this optimum is robust, we subsampled 100 IOS from SMD, ran an all-vs-all pairwise comparison (excluding identical pairs and pairs of different jaw types). This subsampling was done 5 times with replacement, with each subsample constituting a batch. This resulted in 4,894 to 5,074 pairs being included in each batch, with the number of matches ranging from 42 to 44. Figure 2 describes the difference between the matching and mismatch distributions in each batch, measured in Wasserstein distance, as a function of number of keypoints used for the scoring scheme. The higher Wasserstein distance, the better, since it indicates larger difference between the distributions. It is evident that the more keypoints are included, the more non-matching keypoints are included, causing matches and mismatches to become more alike. On the other hand, including fewer keypoints would mean relying on less of the dentition surface to determine the score. In the extreme case where only one keypoint is used for the scoring scheme, a comparison would be considered a match if as little as an area within a 2 mm radius had similar curvatures on the compared IOS. Therefore, there is a trade-off between differentiating between the scoring distributions and include enough keypoints for a robust method. Due to this diminishing return when increasing the number of keypoints included in the analysis, the optimal number of keypoints was set when the mean curve of all five batches has a slope of 75% of the maximum slope, representing the shoulder of the curve. In Figure 2 we see that the optimal number of keypoints to use depends on the scoring metric, with the optimal number of keypoints ranging from 8 to 16.

**Figure 2.**
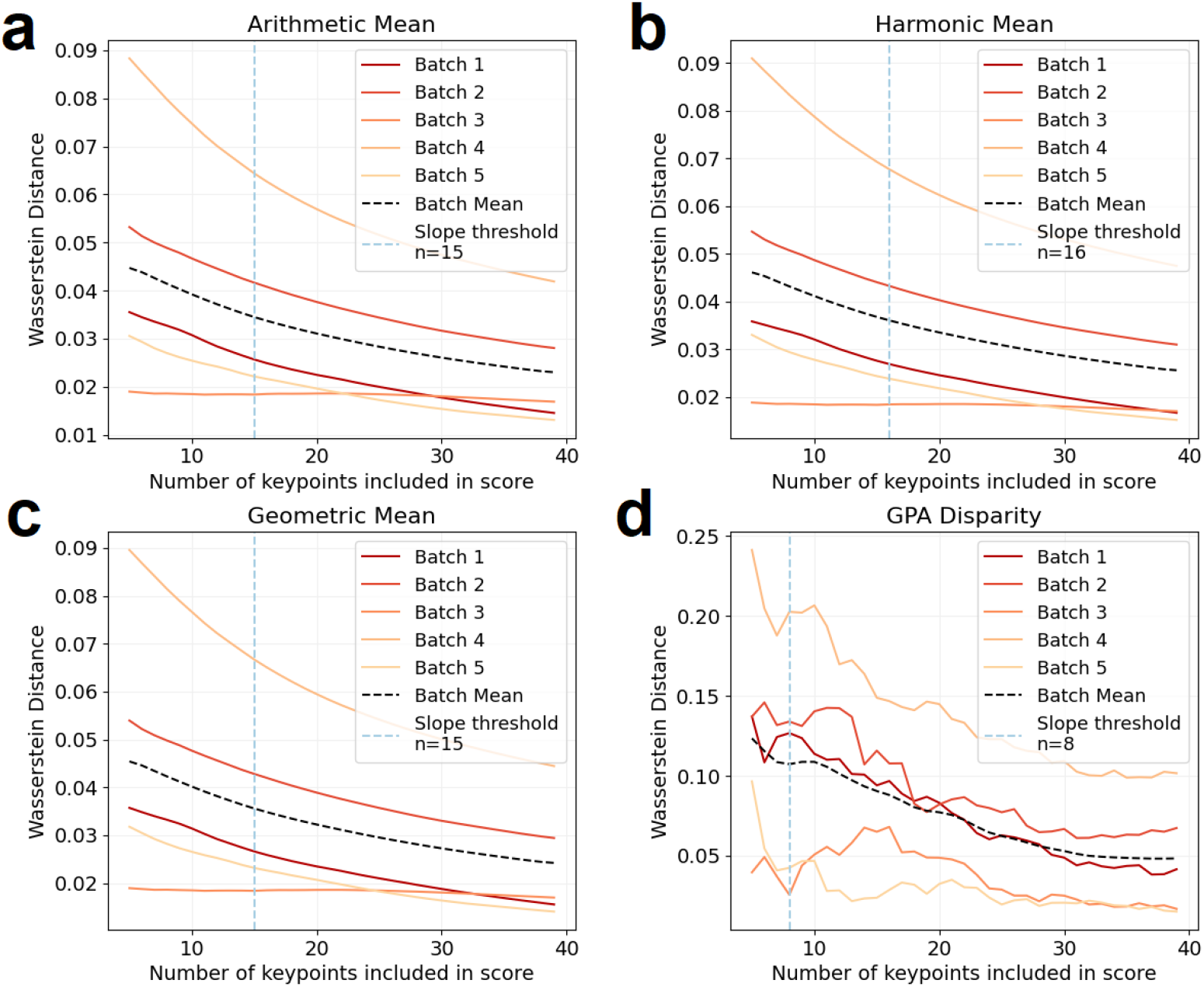
Wasserstein distance of SMD batches with varying number of keypoints. The overlap between matching and mismatching distributions of SMD batches (subsamples) is illustrated as Wasserstein distance between the matches and the mismatches, plotted against the number of keypoints used in the scoring scheme a) arithmetic mean b) harmonic mean c) geometric mean and d) GPA disparity.

To take advantage of both the relative keypoint placement and the surface curvature similarity, the weighted average of scoring schemes was explored. Here, we aim at minimising the overlap between the matching and mismatching distribution, to minimise ambiguity of the resulting score and decrease the likelihood of both false positives and false negatives. This was done by measuring the range of overlap between matches and mismatches when increasing the weight on GPA disparity. In Figure 3 we see the best separability of matches and mismatches in the SMD batches when giving disparity 10% weight and the mean scoring metrics 90% weight. All three mean metrics score similarly, so for the sake of simplicity, the arithmetic mean combined with GPA disparity was chosen as the favoured scoring metric for IOS keypoint comparison. Since the performance increase by using the weighted average is small, the individual scores should also be investigated individually. As discussed previously, since subsets of SMD were used to optimise the scoring scheme, SMD cannot be used to validate its performance without risking performance bias. Therefore, the scoring scheme was validated on the independent HVD dataset.

**Figure 3.**
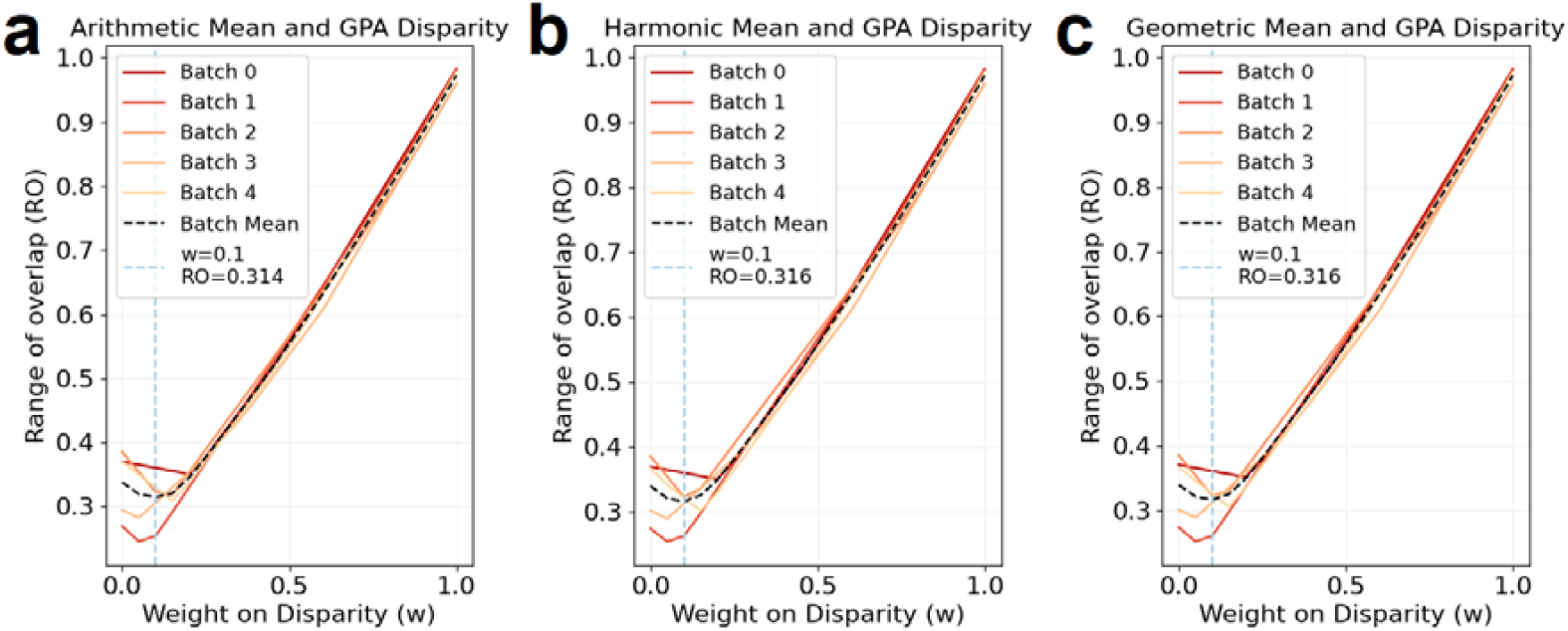
Range of overlap of SMD batches with varying weight on disparity. The overlap between matching and mismatching distributions of SMD batches is illustrated as the range of overlap between the matches and the mismatches, plotted against the weight put on GPA disparity when scoring with a) arithmetic mean and GPA disparity b) harmonic mean and GPA disparity and c) geometric mean and GPA disparity.

### Evaluation of the pipeline in the HVD data

To evaluate the scoring pipeline, an independent dataset was used, namely HVD (cf. Materials and Methods). For the HVD, we tested the original IQR scoring after limiting soft tissue using grid-cutting [17], alongside the newly developed scoring schemes. Figure 4a graphically shows that it was not possible to unambiguously separate matches from mismatches using IQR but matches still showed a different distribution. The overlap is large resulting in a Wasserstein distance of only 0.03 (0 means that two distributions are identical). Figure 4b-d shows the same analysis but using the arithmetic mean of L2 distance ratios of the best matching 15 keypoints, the GPA disparity of the best matching 8 keypoints, and the weighted average, respectively. We see that these new scoring schemes have a greater separation of matches and mismatches, while utilising more range. By comparing Figure 4b-d we see that the combination of arithmetic mean and GPA disparity slightly outperforms using the arithmetic mean alone and using the IQR scores, since the Wasserstein distance is the largest in this case. Even though using GPA disparity as a standalone score has the greatest Wasserstein distance, the overlap between distributions is too great, increasing the risk of false positives and false negatives. The weighted mean (Figure 4d) therefore remains the favoured scoring scheme. Using the favoured scoring scheme, 94.9% of the matches lie outside of the 5^th^ percentile of the mismatching distribution, suggesting that matching scores lie in a range not typically occupied by mismatching scores. It is also evident that the numerical value of the scoring indicates a level of trustworthiness. A comparison with a particularly low score is a confident match, while there is ambiguity when comparisons score within the overlap of the matching and mismatching distribution. In Figure 4e the predictive power of the different scoring schemes was investigated by their individual ability to distinguish between matches and mismatches across all thresholds using ROC-AUC. ROC-AUC focusses on the true match rate (recall) and the false match rate [22]. Here we see that the new scoring scheme outperforms the original IQR scoring scheme with a ROC-AUC of 0.990 (95% CI 0.988 to 0.992). Such a high ROC-AUC indicates that this scoring scheme is considered good at discriminating between matches and mismatches. It is noteworthy that the weighted average of scores and the standalone arithmetic mean performs equally well according to the ROC-AUC. The PR-AUC showed similar performance and can be seen in Online Resource 1.

**Figure 4.**
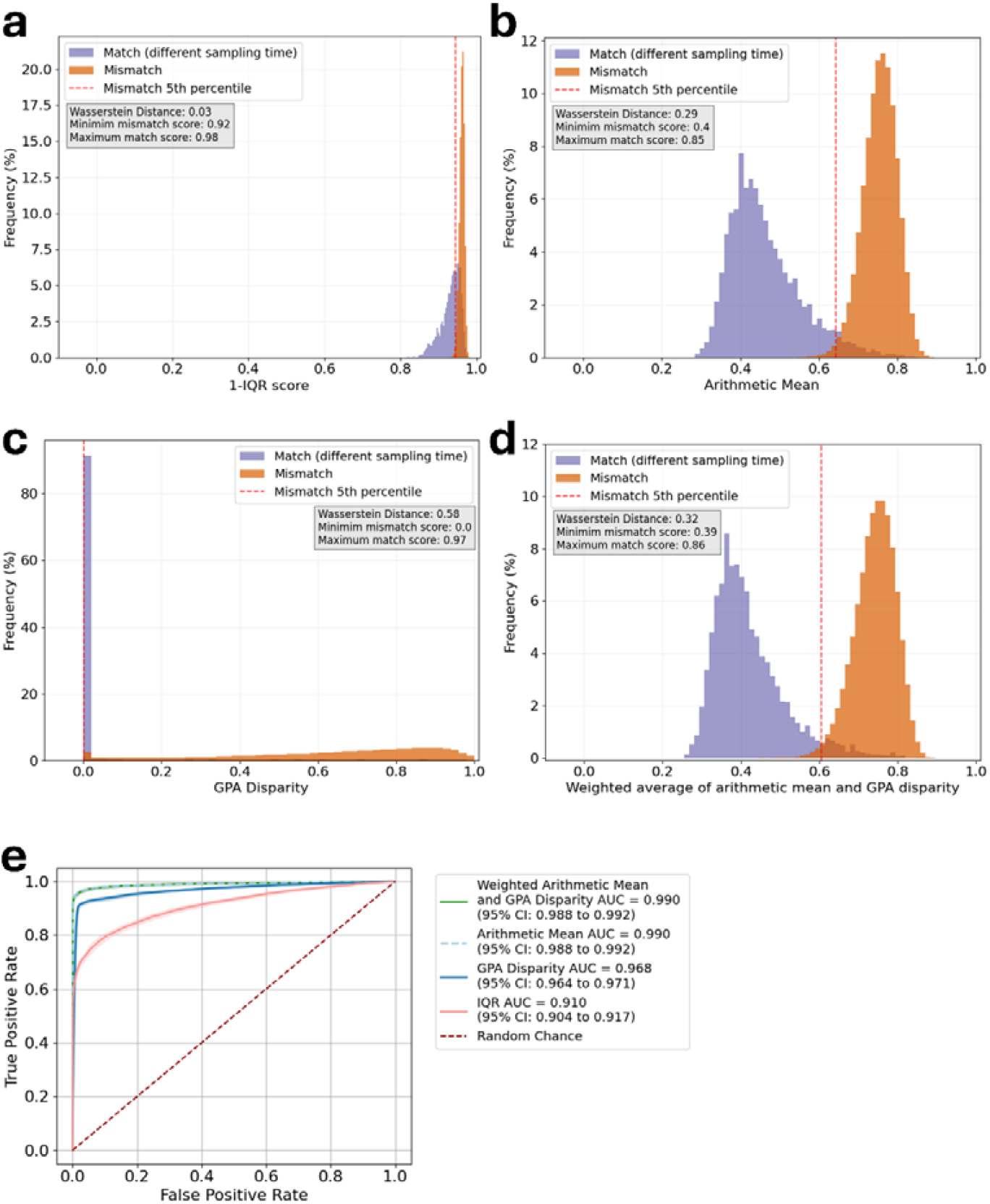
HVD score distributions. Scores for matches and mismatches of IOS comparisons for all IOS included in HVD. The panels show a) distribution based on IQR score, b) distribution based on arithmetic mean of the best matching 15 keypoints, c) distribution based on GPA disparity of the best matching 8 keypoints, d) distribution based on weighted average of arithmetic mean GPA disparity weighted 90%/10% respectively and e) ROC-AUC describing the predictive power of the two scoring schemes.

### Results applied in a forensic setting using ranks

In a forensic setting, the scoring schemes would be used to rank AM dental records according to a most likely match to a specific PM IOS. In such a context, the ranking power of the scoring scheme needs to be explored. In the case of HVD, each IOS can have multiple true matches. This is true since each IOS is included both as a whole dental mesh, and as partial dental meshes. Therefore, in this rank analysis, the rank refers to the number of mismatches that scores better than the particular match. The rank of a true match is 0 if the match is the lowest scoring comparison, or if the match is only outranked by other true matches. As seen in Figure 5, when using the new scoring scheme, 94.49% of the true matches are found at rank 0, and 96.80% of the true matches are found within rank 5. Only 5 cases (0.49%) were considered a false positive case, where a mismatch was the ultimately best scoring comparison. In 4 of those cases, a true match was found within rank 3, and for the last case a true match was found in rank 7. In this case, no true match scored better than 0.63, thus underlining that the numerical value of the similarity score can be used to assess the confidence in the matching. The lowest score given to a mismatch in the particular case was 0.57, showing that even though this mismatch was the best scoring comparison, it would still not have been a confident misclassification, showing that this scoring scheme is robust regarding false positives. This establishes the new scoring scheme as a valid option for ranking AM IOS according to most likely match to a PM IOS.

**Figure 5.**
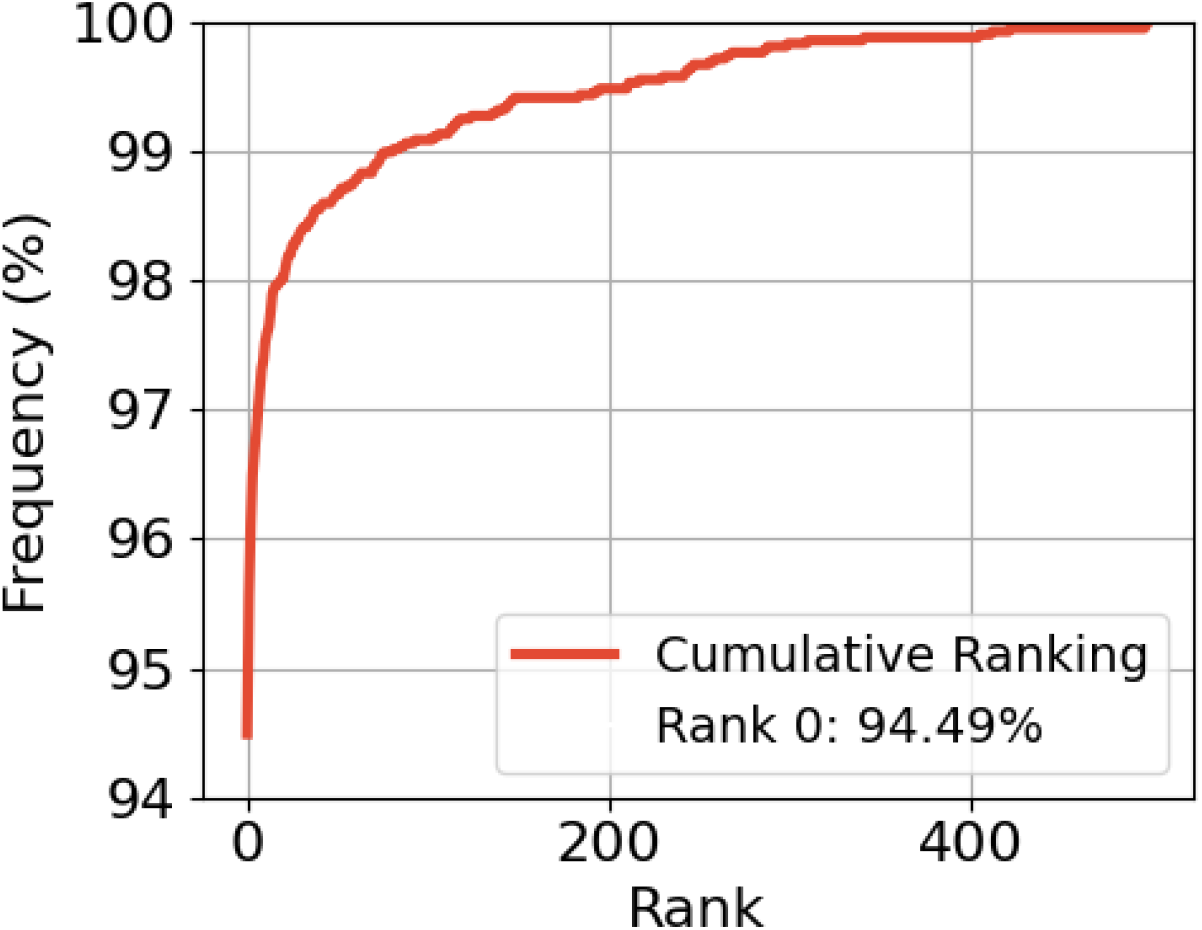
HVD rank distribution. Ranks for true matches of IOS comparisons for all IOS included in HVD. Scoring scheme is weighted average of arithmetic mean of the most similar 15 points and GPA disparity of the most similar 8 points, weighted 95%/5% respectively.

### A close look at the false negative comparisons

We manually investigated the 137 (3.2%) of matching comparisons where more than 5 mismatches score better than the match. We saw that only 10 of these comparisons include a full dental mesh. This means that the false negative cases mainly arise when two partial IOS are compared. In a forensic setting, it would be expected that most AM IOS would be full jaws IOS, thus making such cases less likely in a real DVI scenario.

It is known that using default settings for tissue removal with grid-cutting can cause problems on some IOS [17]. Therefore, the comparisons were checked for excessive surface removal, to evaluate if it was a true scoring problem, or if data preprocessing could have removed parts of the matching dental surface. In 64 of the 137 cases, the bad score could be attributed to a limited matching dental surface between IOS due to removal of dental surface during grid-cutting. In these cases, the scoring is not expected to be able to recognise the match. Removing these comparisons from the analysis, only 1.7% of the matching comparisons had 5 or more mismatches with better scores.

In 123 of the false negative comparisons, partial dental meshes were compared where only a quarter of the full dental surface was a true match, still with 6 months between the scans. In these cases, the score can be inflated if there is not 15 keypoints that are matching, within such a small surface area. One example can be seen in Figure 6a-c. In this example where only a quarter of the jaw matches, the 15 best matching keypoint pairs have been marked. We see that for the 15 best matching keypoints, also keypoints outside of the true matching surface is included, causing mismatching keypoint pairs to be included in the final score. This is expected to increase the score, resultantly decreasing the confidence in the match. Upon evaluation, this phenomenon seems to cause 73 false negatives out of a total of 247,248 comparisons where partial dental meshes are compared with only a quarter of the dental surface being a match.

**Figure 6.**
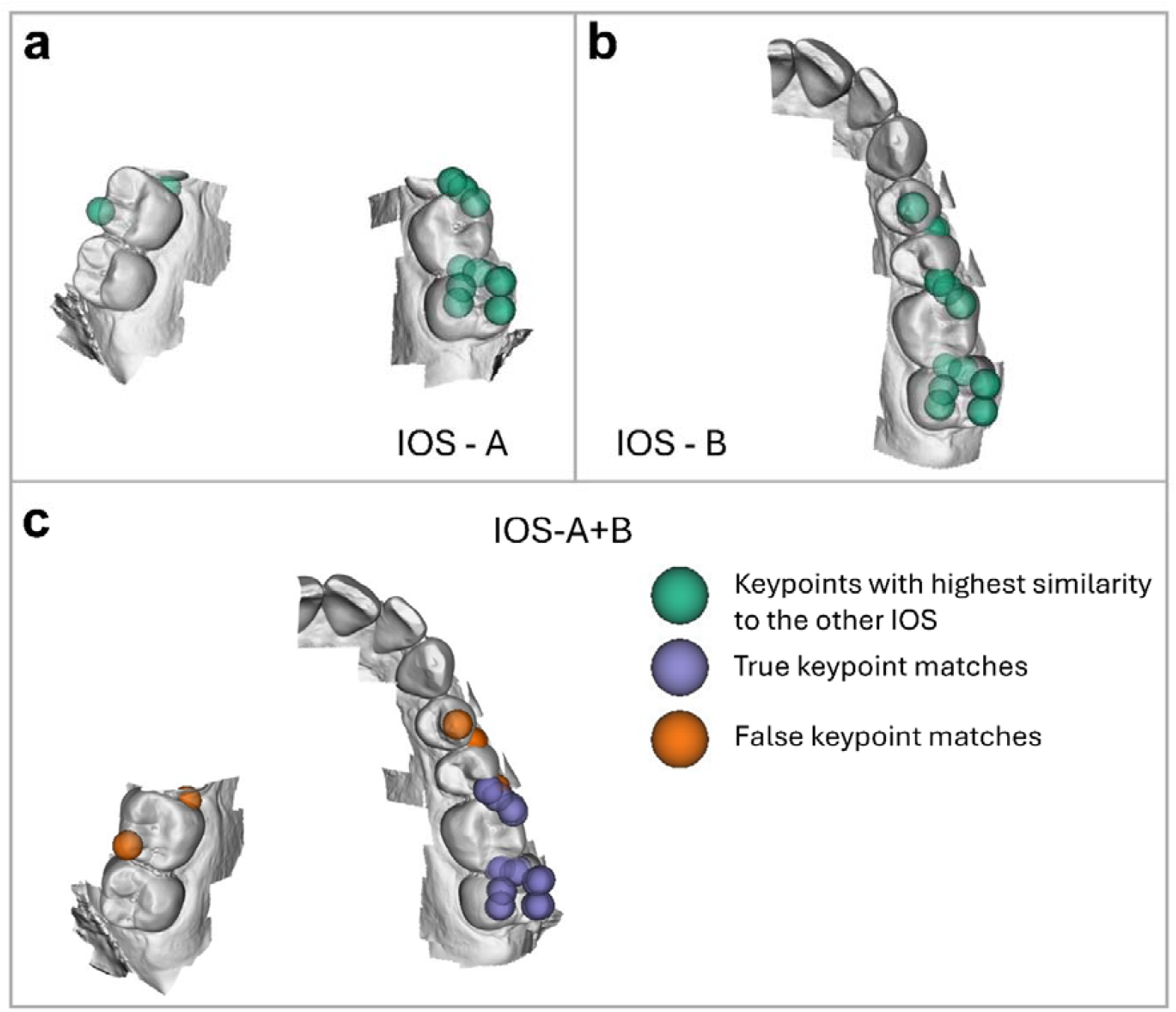
Visualisation of matching instance where only a quarter of the dental surface matches, causing the score to include false matching keypoints (orange), since no more true matching (purple) keypoints are available for such a limited surface. Resultantly, the comparison was scored as a false mismatch. Shown is the keypoints for a) IOS-A, b) IOS-B and c) the overlay of IOS-A and IOS-B.

## Discussion and conclusion

In this study we used IOS data from living individuals gathered at two timepoints to improve the previously proposed pipeline for quantitative scoring of dental surface matching [INREV]. By optimising on the scoring scheme used for differentiation between matches and mismatches, we were able to improve the ability to predict matching IOS. Even though the standalone arithmetic mean had the same ROC-AU as the weighted average between the arithmetic mean and the GPA disparity, the weighted average showed a larger Wasserstein distance, favouring this scoring scheme. The best tested scoring scheme was therefore the weighted average between the arithmetic mean of the 15 most similar keypoints, and the GPA disparity of the 8 most similar keypoints, weighted 90%/10% respectively. This scoring scheme does not include large scale global information, since the keypoint similarity is measured based on representations of the vertices within a 2 mm radius of each keypoint. Therefore, this method should be robust to many intrapersonal changes such as restorative treatment, attrition, or losing one or more teeth. By including the relative placement of the keypoints in the form of GPA disparity, another level of information, the regional information, was added to the scoring scheme. We see that for the dataset used in this study, this combination of information improves separability of the two classes, compared to the previous IQR score [5,22,23]. In this regard, one must consider that AUC shows how well match instances separate relative to mismatch instances and does not show how well the scoring scheme is calibrated to the 0 to 1 scale range [22]. This means that even though the new scoring scheme shows the best separation, it is not guaranteed that this scoring scheme has an intuitive use of the 0 to 1 scale range [22].

Looking further into the individual scoring schemes, we see that the arithmetic mean is better at containing the matching and mismatching distributions within a limited range, while the GPA disparity is better at maximising the mean difference between the distributions. Even though it is desirable to have a large mean difference between matches and mismatches, GPA disparity shows long tails on the distributions, causing a greater overlap between matches and mismatches, which is undesirable. We also see that with a smaller surface overlap between a matching pair of IOS, the fixed number of points used for score calculations might falsely inflate the score, resultantly lowering the confidence in the match. This could possibly be alleviated by using fewer keypoint pairs for the scoring, which would lower the confidence in the scoring. This trade-off between number of keypoints and confidence in scoring underlines that the fewer teeth used for comparing two jaws, the less trustworthy is the match.

We believe that this study utilizes the strengths of using local surface signatures to predict a global label, in this case, whether two IOS comes from the same individual. It does so by focussing on keypoints at extreme dental curvatures and comparing a representation of the local surface curvatures. Furthermore, this study expands the field of 3D dental matching by incorporating previously unexplored measures of local dental surface curvatures, thereby reducing the impact of intrapersonal variation and systematic variation in IOS across long distances [5,14,15].

This study presents easy to read quantitative scoring schemes, that would allow a forensic odontologist to quickly assess whether two IOS are deemed a likely match or an unlikely match. Such quantitative scoring schemes can be used to rank AM databases according to a most likely match, allowing for a quick sorting before identifying victims of major disasters.

Our results showed that the scoring scheme is robust regarding false positives and that 46.7% of the false negatives could be attributed to data preprocessing issues. To alleviate this, one should consider automated removal of soft tissue for data preprocessing followed by manual quality control [17]. Even though the score calculation relies on a fixed number of keypoints, the pipeline showed great discriminatory power, even when as little as a quarter of the dentition surface was matching.

The fact that a fixed number of keypoints are needed for calculating the score, makes the method dependent of an appropriate amount of dentition surface to be included in the analysis. As shown, this only seldomly constitutes an issue, underlining the robustness of the developed pipeline and scoring scheme.

## Supporting information

Online Resource 1

## Data Availability

The research data consists of GDPR-protected data that the authors are not authorized to share. The code used in this study can be found here: https://github.com/AnikaKofodPetersen/Scoring_scheme

## Statements and Declarations

### Ethics Approval

The study was carried out in adherence to ethical principles, including the guidelines outlined in the World Medical Association’s Declaration of Helsinki. The study is registered with the Data Protection Unit at Aarhus University (file number 2016-051-000001, serial number 2534; file number 20220367531, serial number 3155). The study and its protocol were initially submitted to the Central Denmark Region Committees on

Health Research Ethics. This committee referred to The Danish National Committee on Health Research Ethics (NVK). NVK assessed that the study and its protocol were exempt from notification since the study does not aim to generate new knowledge about the origins, prevention, diagnosis, or treatment of diseases (case number: 2400741). Prior to acquisition of SMD, the study protocol and data retrieval protocol were additionally approved by the Regional Council and the Silkeborg Municipality Council (case number: 1-45-70-49-2; case number: EMN-2023-02524).

### Consent to participate

Written informed consent was obtained from all individuals contributing to the HVD data set. The SMD dataset was exempted from this regulation, decided by the Regional Council and the Silkeborg Municipality Council.

### Competing Interests

The authors have no relevant financial or non-financial interests to disclose. The authors have no competing interests to declare that are relevant to the content of this article.

The authors affirm that human research participants provided informed consent for publication of the images in Figure 1 and Figure 6.

### Funding

This study was funded by AUFF NOVA (Grant number: AUFF-E-2021 -9-14).

## Notes

### Competing Interest Statement

The authors have declared no competing interest.

### Author Declarations

The Danish National Committee on Health Research Ethics declared exception of notification for this work

