## Supplementary figures and images for "Objective comparison of 3D dental scans in forensic odontology identification"

### Online Resource 1

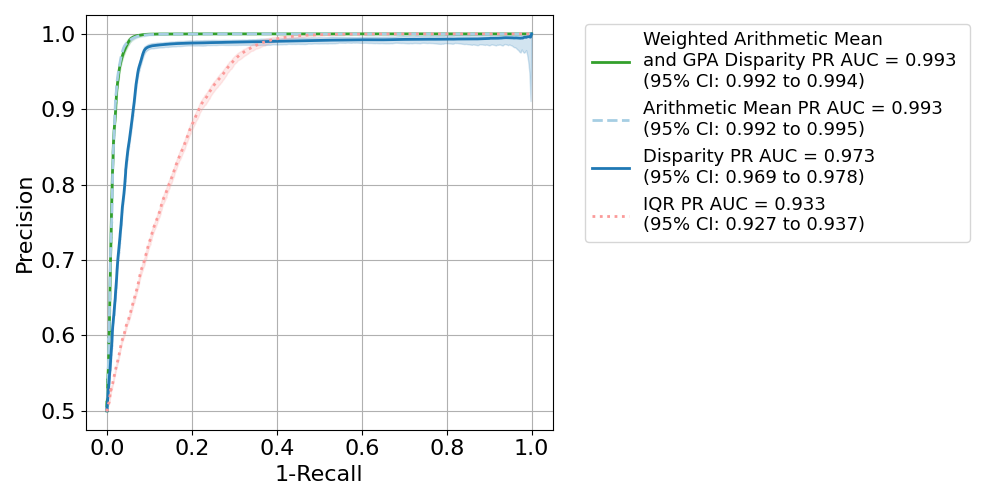
